# Evaluating Nursing Performance During Heatwave Simulations: Protocol for a Repeated Measures Cross-Over Study

**DOI:** 10.64898/2026.09.18.26363404

**Authors:** Hannah Blount, Leisa Bingham, Nuno Koch Esteves, Thomas Daniels, Victoria Filingeri, Carlos Aceves-Gonzalez, Katya Brooks, Natasha Watts, David Jones, Adam Tewksbury, Katie Jenkins, Patrick James, Tracy Sach, Ana Raquel Nunes, Tom Roberts, Chiara Dall’Ora, Mari Carmen Portillo, Chloe Maguire, Liz Raisbeck, Tommaso Costantini, Paul Clarkson, Paul West, Nicky Street, Ralph Gordon, Peter R Worsley, Davide Filingeri

## Abstract

This study protocol combines clinical simulations with climatic chamber experiments, to evaluate 1) the impact of heat stress on hospital nurses’ physiological strain, physical and mental fatigue, vigilance, and clinical performance; and 2) the efficacy of a practical cooling intervention (i.e. electric fan) in mitigating the negative impact of heat stress.

UK-based, registered adult nurses working in hospital settings will be recruited to partake in a repeated-measures cross-over design study, consisting of five separate 3-hour experimental sessions simulating clinical work, and taking place within a climatic chamber at Southampton General Hospital (UK). Sessions will differ by ambient temperature and cooling intervention as follows: 1) control (CTR; 18°C, 50% relative humidity [RH]); 2) warm (WARM; 26°C, 50%RH); 3) warm with electric fan (WARM+FAN; 26°C, 50%RH); 4) hot (HOT; 34°C, 50%RH); 5) hot with electric fan (HOT+FAN; 34°C, 50%RH). This study has been registered on a publicly accessible clinical trial registry (ISRCTN45793399).

During each session, participants will first complete a standardised fatigue-induction protocol simulating shift work (i.e. low-intensity cycling with cognitive tasks); followed by a simulated hospital ward round involving six simulated patients and two simulated cardiac arrest scenarios. Nursing cognitive and physical performance will be assessed using a standardised observational assessment battery, along with objective cardiopulmonary resuscitation (CPR) performance metrics. Physiological responses will be assessed by monitoring changes in core temperature (T_core_), skin temperature (T_skin_), heart rate, and whole-body sweat loss (WBSL). Fatigue and vigilance will be measured using the Swedish Occupational Fatigue Inventory (SOFI) and Brief Psychomotor Vigilance Task (PVT-B), respectively.

Baseline (i.e. prior to fatiguing protocol) to post simulation changes in 1) vigilance assessed via the PVT-B (i.e. a determinant of role-specific cognitive performance); 2) CPR quality score (i.e. a determinant of role-specific physical performance); and 3) T_core_ (i.e. a determinant of thermo-physiological state); will be analysed using linear mixed models to determine the effects of ambient temperature and cooling intervention on nurses’ clinical and physiological performance.

By providing a controlled methodological framework for assessing the effects of heat exposure on hospital nurses’ physiological, perceptual, and clinical performance, this study will provide empirical evidence on air temperature-related changes in nursing performance and physiological strain and the role of cooling interventions in modifying these responses. This will support the development of evidence-based climate adaptation strategies to protect healthcare staff wellbeing and maintain patient safety during periods of extreme heat.

## 1. Introduction

Climate change, and the resulting increase in extreme weather events such as heatwaves, is reshaping the operating environment for healthcare providers worldwide (1, 2). In the United Kingdom (UK), the increase in frequency and intensity of recent heatwaves (3–5) has led the Climate Change Committee to recognise heat as the top environmental hazard facing the country and its healthcare systems (6).

Since ∼2020, hospitals within the UK National Health Service (NHS), many of which lack adequate cooling infrastructure (7), have experienced a 50% rise in overheating incidents (8). Air conditioning in most UK hospitals remains limited to operating theatres, intensive care units, and some specialist areas (7), largely due to sustainability challenges associated with high carbon footprint, aging infrastructure, energy demands, and costs (9).

The health consequences of extreme heat for vulnerable patient populations are well established, including increased risks of morbidity and mortality (4, 10), with climate-driven infectious disease patterns adding operational strain (11). However, heat exposure also presents significant challenges for healthcare professionals such as nurses, who are responsible for delivering safe and effective care during periods of elevated demand resulting from heatwaves.

Healthcare professionals frequently experience prolonged occupational heat exposure while undertaking physically and cognitively demanding clinical tasks (12, 13). Extreme heat is of particular concern within healthcare settings due to its negative impact on fatigue (14, 15) and productivity (16), both of which have been recently identified to contribute directly and indirectly to patient harm by the Health Services Safety Investigations Body (17). These risks are further compounded by existing healthcare workforce pressures, including staffing shortages and increasing service demand during heatwaves (18, 19).

Despite growing recognition of heat as a threat to healthcare systems, important evidence gaps remain (1). Current guidance, including the UK Health Security Agency Adverse Weather and Health Plan for England (20) and the NHS Climate Adaptation Framework (21) for organisations in England, provides recommendations for managing heat-related risks and strengthening organisational preparedness, including consideration of staff vulnerabilities during extreme weather (21). Yet, implementation across healthcare settings remains inconsistent, due to insufficient evidence on 1) the ambient temperature conditions at which healthcare professionals’ clinical performance and patient care quality may begin to decline; and 2) the system impact and cost-effectiveness of different heat adaptation strategies (e.g. cooling) specifically addressing the health and wellbeing of staff such as hospital nurses (22).

Studying healthcare professionals’ performance in real hospital settings, and during naturally occurring heatwaves, presents methodological and logistical challenges associated with variability in environmental conditions, patient needs, staffing levels, and organisational contexts (23). Clinical simulations have a long history in healthcare education (24), and have recently gained recognition as a promising research technique for supporting improvement in healthcare systems and processes (25). Similarly, climatic chamber experiments have been long used to impose controlled heat stress, and to study humans’ physiological and perceptual responses that are relevant to a variety of occupations (e.g. outdoor workers, military personnel, fire brigades, etc. (26)). Combining these approaches could therefore offer a safe and reproducible alternative to investigating the physiological, cognitive, and clinical implications of heat exposure on staff performance under live clinical settings. Such simulations could also provide an opportunity to perform initial evaluations of infrastructural and user-centred cooling strategies, including active cooling (e.g. to reduce ward temperatures) and lower-energy ventilation approaches (such as electric fan-assisted cooling), to mitigate heat strain during clinical work, which could inform local heatwave plans (e.g. trigger air temperatures for implementation of interventions) as well as future revisions of the Adverse Weather and Health Plan for England. A simulation-based approach has been recently and successfully applied in other occupational settings (e.g. ready-made garment factory industry (27)) to inform economic modelling of potential cost-benefits associated with selected interventions, which can in turn drive policy change and investment within the UK healthcare sector (6).

It is within the context above that this study protocol aims to combine clinical simulations with climatic chamber experiments, to evaluate 1) the impact of heat stress on hospital adult nurses’ physiological strain, physical and mental fatigue, vigilance, and clinical performance; and 2) the efficacy of a practical cooling intervention (i.e. electric fan) in mitigating the negative impact of heat stress.

By providing a controlled methodological framework for assessing the effects of heat exposure on hospital nurses’ physiological, perceptual, and clinical performance, this study will provide empirical evidence on air temperature-related changes in nursing performance and physiological strain and the role of cooling interventions in modifying these responses. These findings will support future evaluation work within live clinical settings, which will ultimately inform national and local climate-adaptation strategies to protect healthcare professionals’ wellbeing and maintain patient safety during periods of extreme heat.

## 2. Materials and Methods

### 2.1 Overview

UK-based, registered adult nurses working in UK hospital settings will be recruited to partake in a repeated-measures cross-over design study, consisting of five separate 3-hour experimental sessions simulating clinical work, and taking place within a climatic chamber at Southampton General Hospital (UK).

Sessions will differ by ambient temperature and cooling intervention as follows: 1) control (CTR; 18°C, 50%RH); 2) warm (WARM; 26°C, 50%RH); 3) warm with electric fan (WARM+FAN; 26°C, 50%RH); 4) hot (HOT; 34°C, 50%RH); 5) hot with electric fan (HOT+FAN; 34°C, 50%RH). Sessions will be performed in a pseudo-randomised order and will be separated by a minimum of 48h, to minimise potential order and learning effects.

The choice of ambient temperature conditions was driven by clinically meaningful considerations and practice within the UK. Ambient temperature for the CTR session was derived based on the design conditions set in the “NHS Health Technical Memorandum 03-01 Specialised ventilation for healthcare premises Part A” (28), whereby general wards are expected to be maintained at 18°C. Ambient temperature for the WARM session was derived based on guidance set by the “UKHSA Heat-Health Alert action card for health and social care providers”, which states that rooms and cool areas should be maintained below 26°C, thereby considering this temperature as a first heat threshold (29). Ambient temperature for the HOT session was derived from overheating incident reports collected at two NHS hospitals in the south of England, which confirmed clinical areas to have reached indoor temperatures of up to 34°C during the 2022 UK heatwave (30). FAN conditions were selected based on the use of a pedestal, electric fan generating an air speed of 3.5m/s (31).

### 2.2 Participants

Participants will consist of UK-based, registered adult nurses working in UK hospital settings, and will be recruited according to the criteria presented in Table 1. The characteristics of the majority of current nurses within the UK workforce are as follow: typically a woman, with an average age of 44 years, practicing adult nursing, and with an ethnicity ratio of 3:1 between white British and other ethnic minorities (e.g. Black, African, Caribbean, or Asian British/non-British workers (32)). We appreciate that our sample will provide a healthy cohort of nurses who may be healthier than the average nursing population. Within this context, should any adverse physiological or performance effects be observed in our cohort, we expect these effects to be amplified in nurses with greater heat susceptibility, such as those with higher BMI, pre-existing health conditions, or altered thermoregulation. The schedule of participant enrolment, interventions, and assessments can be found in Figure 1.

**Figure 1.**
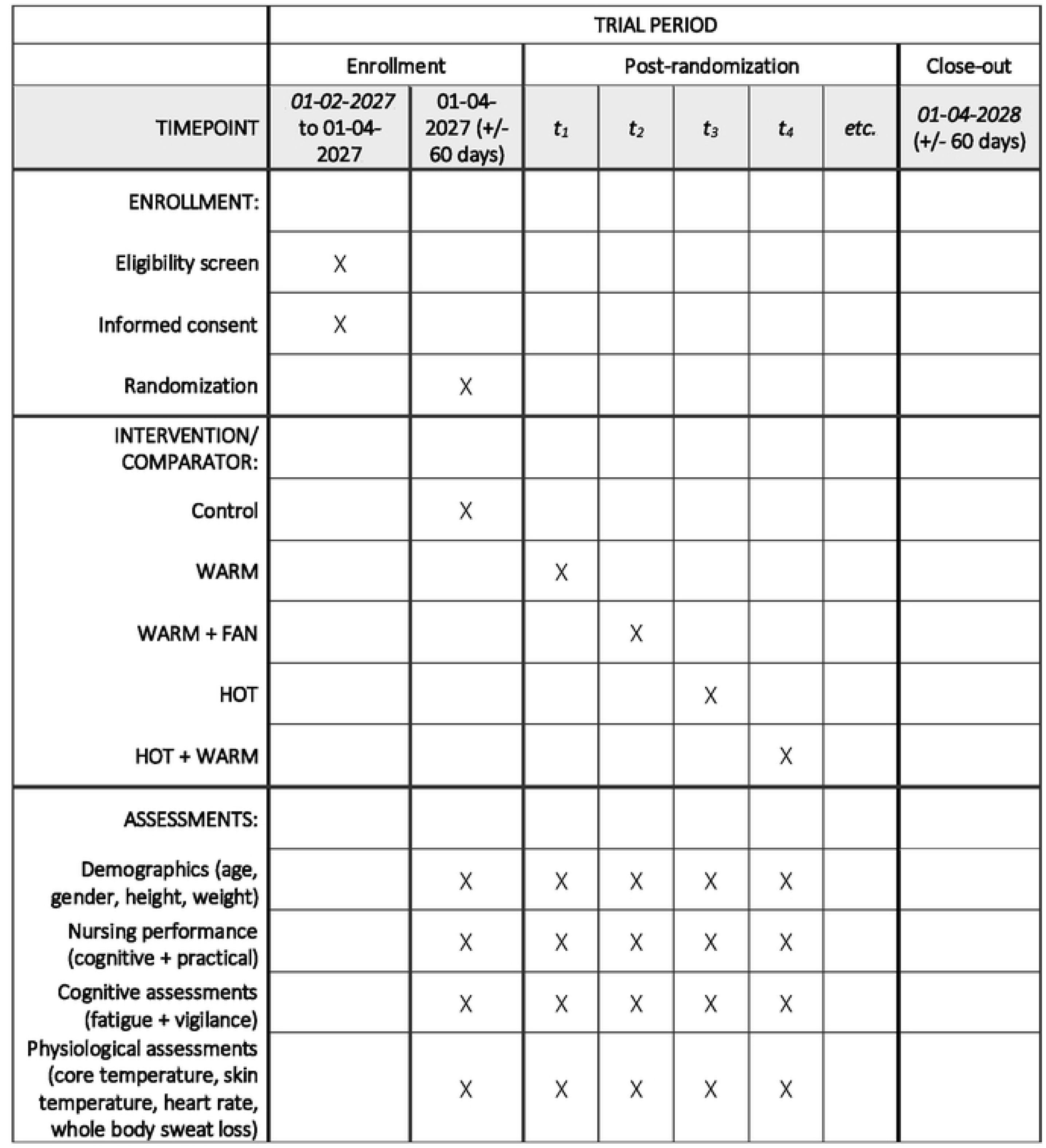
SPIRIT participant timeline (56).

**Table 1.** Participant inclusion and exclusion criteria.

| Inclusion | Exclusion |
| --- | --- |
| 1. Capable of providing informed consent | 1. Female participant is pregnant |
| 2. Willing and able to attend all visits | 2. History of cardiovascular disease |
| 3. Healthy male or female volunteers | 3. Suffering from neurological conditions |
| 4. Aged 18-65 years | 4. Suffering from skin conditions (e.g. eczema) |
| 5. Physically active (i.e. performing 30 minutes regular exercise of moderate intensity, physical activity on at least 3 days each week for at least 3 months) | 5. Presence of conditions altering thermoregulation and blood flow (e.g. Raynaud’s disease) |
| 6. Registered nurse | 6. Smoker or vaper |
|  | 7. Allergic to adhesives |
|  | 8. History of gastrointestinal obstruction, dysphagia, swallowing difficulties |

### 2.3 Experimental Procedures

Once screened and recruited, participants will be invited to complete five experimental sessions at Southampton General Hospital (UK) on separate days.

Participants will ingest a telemetric temperature capsule (e-Celsius Performance pill, BodyCAP, France) approximately 3–4 h before each experimental session for continuous T_core_ measurement. Upon arrival, height (Seca 213 Stadiometer, Birmingham, UK), body mass (KERN 150K2DL, Balingen, Germany; accurate to 0.005 kg), and demographic information will be recorded during the initial visit. For the assessment of WBSL, dry nude body mass will be measured before each trial behind a privacy curtain, and participants will be instructed to refrain from drinking until the post-trial body mass measurement has been completed. For all experimental visits, participants will be instrumented with a heart rate monitor (POLAR H10, POLAR Electro, Kempele, Finland) and four wireless T_skin_ sensors (iButtons, Maxim, San Jose, USA; 0.2Hz) positioned on the calf, thigh, chest, and shoulder such that mean T_skin_ can be calculated with established equations (33). Participants will wear their standardised nursing uniform which will be the same for each experimental visit.

Following instrumentation, participants will enter the climatic chamber and undergo an initial assessment of fatigue and vigilance. Subjective fatigue will be assessed using the Swedish Occupational Fatigue Inventory (SOFI) (34, 35), whilst objective vigilance will be assessed using the Brief Psychomotor Vigilance Task (PVT-B) (36), for which, the software PsyToolkit will be used (37, 38). Participants will also perform a baseline assessment (i.e. pre-fatigue) of cardiopulmonary resuscitation (CPR) on a manikin, and they will not be informed of the required duration, replicating the uncertainty of real-world emergencies. They will be instructed to continue until stopped by a researcher; however, to ensure consistent physical requirements across participants, each CPR bout will be standardised to 2 min. CPR performance will be quantified using software integrated within a resuscitation manikin (Resusci Anne QCPR, Laerdal Medical, Kent, UK).

Participants will then complete a standardised fatigue-induction protocol consisting of 30 min of low-intensity cycling on a semi-recumbent ergometer (∼20 W; ∼2 METs; 60 rpm). During cycling, participants will simultaneously perform a cognitive battery comprising three Key Stage 2 arithmetic tasks interspersed with a Stroop Colour and Word Test. This protocol has been designed to approximate light physical activity which equates to the average energy expenditure during a nursing shift (i.e. ∼2 METs) (39), while also incorporating cognitive tasks previously shown to induce mental fatigue and reduce reaction times (40). Immediately following the fatigue-induction protocol, participants will repeat the SOFI and PVT-B assessments to quantify changes in fatigue and vigilance prior to commencing the nursing simulation.

At this point, participants will begin a simulated clinical shift lasting approximately 105 min. The climatic chamber will be configured to replicate a side-room hospital ward environment (Figure 2). Participants will receive a standardised handover consisting of six simulated patients and will subsequently complete a sequence of routine nursing activities. Tasks will include patient communication, vital-sign assessment and recording, medication management, recall of handover information, and infection-prevention procedures. A trained research team member will portray the 6 simulated patients using standardised scenarios. During the simulated ward round, participants will also complete two simulated cardiac arrest scenarios, occurring after the second and fourth patient encounters. In each scenario, participants will initiate CPR on a manikin for a duration unknown to them and will be instructed to continue until stopped by a researcher. To ensure consistent physical demands across participants and conditions, each CPR bout will be standardised to 2 min.

**Figure 2.**
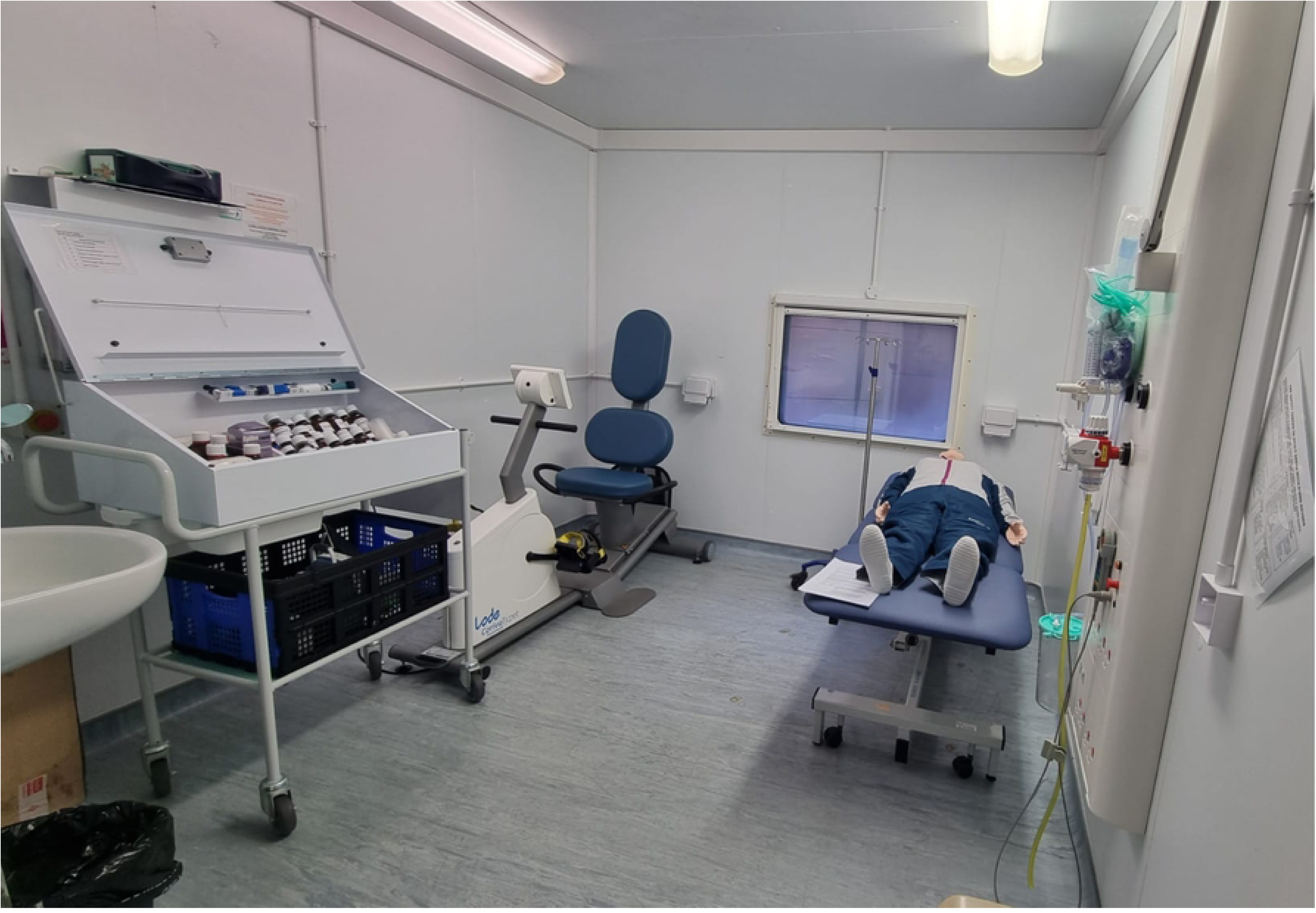
Climatic chamber set up with a medication trolley, hospital bed with a CPR manikin and an exercise bike.

Clinical performance will be assessed throughout the simulation using a standardised observational assessment battery completed by a member of the research team (available in the supplementary material). Assessment domains will include communication, consent, hand hygiene, personal protective equipment use, vital-sign assessment, medication management, and CPR performance. The assessment battery was developed from existing simulation-based education and assessment practices within a higher education adult nursing programme, and reflects domains commonly assessed within Objective Structured Clinical Examination (OSCE)-style approaches to nursing education and clinical skills assessment, including communication, procedural accuracy, medication safety, infection-prevention behaviours, and clinical decision-making (41). This approach is also consistent with professional support for simulation-based learning in nursing education, including the Royal College of Nursing (42) and Nursing & Midwifery Council (43) guidance recognising simulation as a method for developing and assessing practice-relevant knowledge, behaviours and skills.

Following the simulation, participants will repeat the SOFI and PVT-B assessments, and dry nude body mass will be measured again for calculation of WBSL. T_core_, T_skin_, heart rate, and environmental conditions will be monitored continuously throughout testing. Upon completion of all assessments, participants will exit the climatic chamber and remain under observation until physiological indicators confirm recovery from heat exposure. An outline of the study protocol can be found in **Figure 3**.

**Figure 3.**
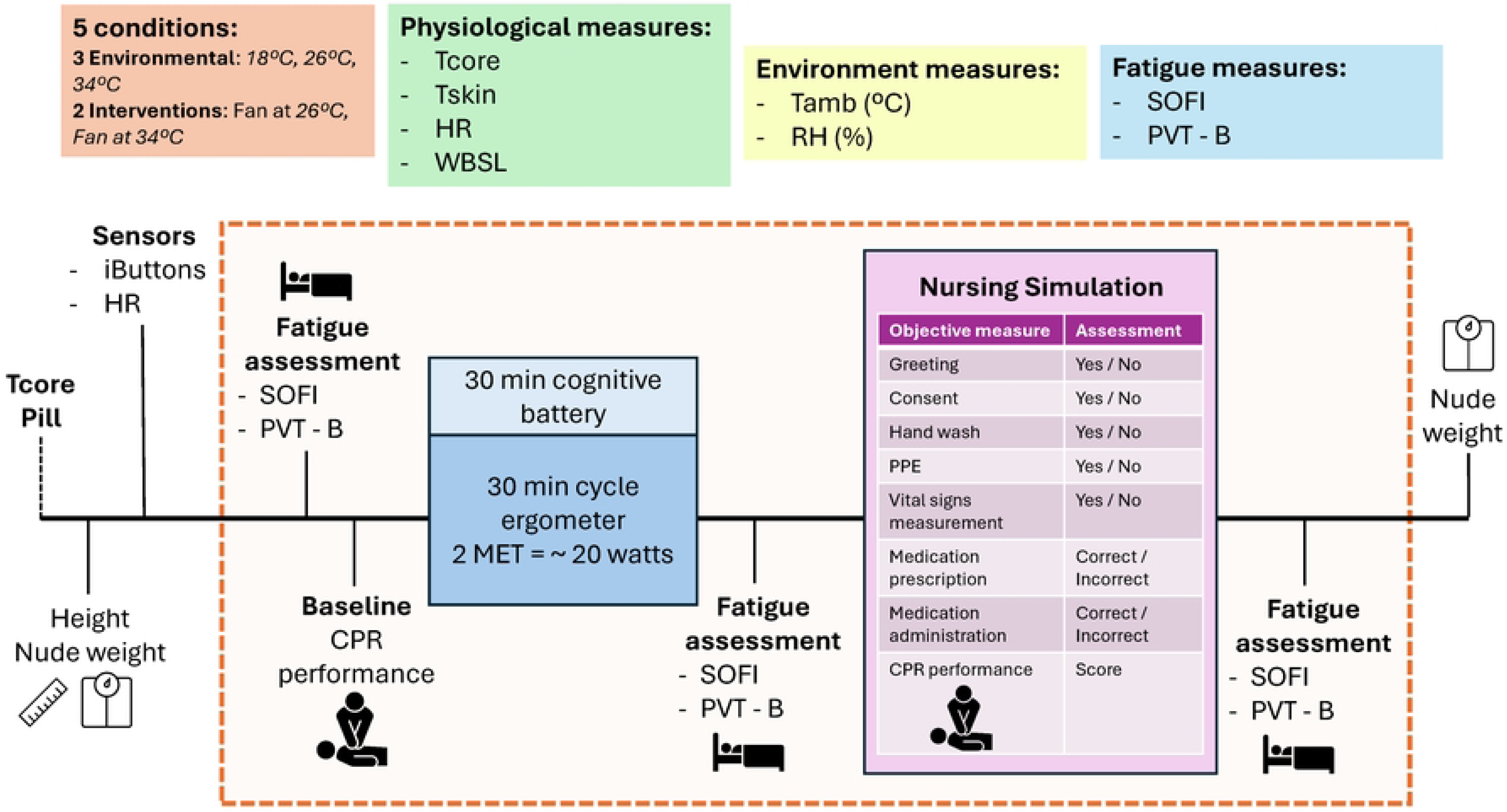
Protocol schematic. Dashed box highlights the elements of the protocol conducted within the climatic chamber. T_core_ = core temperature. T_skin_ = skin temperature. HR = heart rate. WBSL = whole body sweat loss. T_amb_ = ambient temperature. RH = relative humidity. SOFI = Swedish Occupational Fatigue Inventory. PVT-B = Brief Psychomotor Vigilance Task.

### 2.4 Measurements

#### 2.4.1 Nursing performance: role-specific cognitive tasks

Clinical performance will be assessed throughout the simulated nursing shift using a standardised observational assessment battery completed by a member of the research team; the full assessment battery is provided in the supplementary information. Participants will complete six standardised patient encounters, portrayed by a single trained research team member using consistent scenarios across conditions. For each patient encounter, completed behaviours will be scored as binary outcomes, and accuracy-based behaviours will be scored as correct or incorrect. A total nursing performance score will be calculated as the percentage of correctly completed behaviours across all six patient encounters. These assessments will be used solely as research outcome measures and will not constitute an assessment of participants’ professional competence. Individual performance data will not be shared with employers, managers, colleagues, professional bodies, or any individuals outside the research team. The clinical behaviours assessed will include greeting the patient, gaining consent, hand hygiene, vital-sign measurement, accurate vital-sign recording, appropriate medication-related decision-making, including questioning the medication chart in response to abnormal vital signs, and correct medication administration. These behaviours were selected because they represent safety-critical components of routine nursing care where omissions, inaccuracies, or delayed decisions may indicate fatigue-related deterioration in attention, memory, clinical reasoning, or procedural accuracy (44). Under heat stress, an increase in missed behaviours, recording errors, medication-related errors, or infection-prevention omissions may therefore provide clinically relevant indicators of impaired nursing performance and reduced quality of patient care. This evidence-based simulation approach supports standardised assessment of clinically relevant behaviours that may be vulnerable to thermal strain, fatigue, and cognitive load during occupational heat exposure. To minimise assessor subjectivity, the assessment battery will focus on predefined, directly observable behaviours scored using binary or correct/incorrect criteria. The feasibility of independent secondary review will be explored, and, where available, a random sample of assessments will be reviewed by a second assessor to support inter-rater reliability.

#### 2.4.2 Nursing performance: role-specific physical task

CPR performance will be assessed at baseline (prior to the fatiguing protocol) and during two simulated cardiac arrest scenarios occurring after the second and fourth patient encounters. Participants will be instructed to continue CPR until directed to stop by a researcher but will not be informed in advance of the required duration. Each CPR bout will be standardised to 2 min across all sessions. CPR performance will be quantified using feedback software integrated within the resuscitation manikin (Resusci Anne QCPR, Laerdal Medical, Kent, UK). Derived CPR outcomes from the software will include compression depth, compression rate, hand positioning and overall CPR quality score.

#### 2.4.3 Cognitive assessments: Fatigue and Vigilance

Subjective fatigue will be assessed using the SOFI, and objective vigilance will be assessed using the PVT-B. The SOFI was selected because it is a multidimensional measure of acute work-related fatigue, capturing lack of energy, physical exertion, physical discomfort, lack of motivation, and sleepiness, which are directly relevant to the combined physical and cognitive demands of simulated nursing work under heat stress (34, 45). The PVT-B was selected because the Psychomotor Vigilance Test is a widely used, reliable measure of behavioural alertness and vigilant attention in occupational roles, including nursing, and the brief 3-minute version has been shown to retain sensitivity to fatigue-related vigilance impairment while being more practical for repeated use in applied settings (36, 46). These assessments will be performed at three time points: before the fatigue-induction protocol, immediately after the fatigue-induction protocol, and following completion of the simulated nursing shift. These measures will be used to determine whether thermal exposure and simulated clinical workload result in measurable changes in fatigue and vigilance across experimental conditions.

#### 2.4.4 Physiological assessments: core temperature

T_core_ will be continuously monitored using an ingestible telemetric temperature capsule (e-Celsius Performance pill, BodyCAP, France). Participants will ingest the capsule approximately 3–4 h prior to each experimental session to allow gastrointestinal transit before testing. Participants will be instructed to wear a wristband for the following 72 h to identify that they had swallowed an MRI incompatible device. During the experimental session, data from the pill will be sampled every 30 s to the receiver (EQ-eViewer Performance monitor, BodyCAP, France). T_core_ was selected as the primary physiological outcome because it provides a robust index of whole-body thermophysiological strain during heat exposure and has been used to justify the sample size calculation for the present study (47). T_core_ responses will also be used for safety monitoring, with testing terminated if predefined safety thresholds are exceeded, i.e. T_core_ > 39°C.

#### 2.4.5 Physiological assessments: mean skin temperature

Local T_skin_ will be measured using four wireless T_skin_ sensors (iButtons, Maxim, San Jose, USA; 0.2 Hz), positioned on the calf, thigh, chest, and shoulder. Sensors will be attached to the skin using adhesive tape prior to entry into the climatic chamber and will remain in place for the duration of the experimental protocol. Mean T_skin_ will be calculated from the four measurement sites using an established weighted approach (33). T_skin_ will be used to evaluate the effect of ambient temperature and fan-assisted cooling on peripheral thermal strain, and to support interpretation of dry heat exchange during simulated nursing work (48). Combined measurements of T_core_ and T_skin_ also allow for the calculation of mean body temperature (49). Furthermore, as T_skin_ is also an important determinant of thermal sensation and comfort (50), these measurements will provide physiological context for participants’ perceptual responses to heat exposure and cooling.

#### 2.4.6 Physiological assessments: heart rate

Heart rate will be monitored continuously using a chest-worn heart rate monitor (POLAR H10, POLAR Electro, Kempele, Finland). Heart rate provides a non-invasive index of cardiovascular strain during the heat exposure, low-intensity physical activity, and simulated clinical work. Heart rate responses will also be used for safety monitoring, with testing terminated if predefined safety thresholds are exceeded, i.e. HR > 80% age-predicted HR_max_.

#### 2.4.7 Physiological assessments: whole body sweat loss

WBSL will be calculated from changes in body mass across each experimental session. Upon entry to the climatic chamber, participants will be instructed to fully undress behind a privacy curtain and dry nude body mass will be measured on a precision scale (KERN 150K2DL, Balingen, Germany; accurate to 0.005 kg). Following the body weight measurement, participants will be instructed not to drink throughout the trial until being weighed post-trial. WBSL will be calculated as the difference between pre- and post-trial body mass. WBSL will provide an additional physiological marker of heat strain across the five experimental conditions.

### 2.5 Statistical Analysis

We selected 3 primary outcomes with robust, underlying empirical evidence on effect sizes, namely 1) changes in vigilance assessed via the PVT-B (i.e. a determinant of role-specific cognitive performance) (46); 2) changes in CPR quality score (i.e. a determinant of role-specific physical performance) (51); and changes in T_core_ (i.e. a determinant of thermos-physiological state)(47). These primary outcomes will be used to determine the effects of ambient temperature and cooling intervention on nurses’ clinical and physiological performance. Primary analyses will focus on change from baseline to post-simulation for PVT-B and T_core_, and change from baseline CPR to the mean CPR quality score recorded during the two simulated arrest scenarios.

Sample size calculations were performed using Gpower (Gpower 3.1) for a repeated-measures ANOVA assessing the within-subject effects of condition and time (α = 0.05, power = 0.95, one group, ten repeated measurements representing five environmental conditions and two assessment time points, i.e. pre- and post-fatigue).

A conservative medium effect size (f = 0.30) was selected as the lower-bound estimate across the three primary outcome domains. This was based on published evidence demonstrating changes in 1) PVT-B scores among female shift workers, where reaction times increased by 0.25s from pre-to post-shift corresponding to an approximate effect size of f = 0.39 (46); 2) CPR performance under rescuer fatigue, where the proportion of compressions reaching recommended depth declined from 53% in minute 1 to 38% in minute 5, corresponding to an approximate effect size of f = 0.30 (51); and 3) T_core_ during simulated occupational tasks under heat stress, where Wibowo et al. reported an approximate 0.5°C increase in body temperature between 22°C and 27°C ambient conditions in healthcare workers, corresponding to an approximate effect size of f = 0.30 (47). As these values indicate medium-sized but practically meaningful changes across cognitive performance, physical performance, and physiological strain, f = 0.30 was selected as a conservative common estimate for sample size calculation. Based on these assumptions, the required sample size was estimated to be 15 participants. Participants will be recruited according to the criteria in **Error! Reference source not found.**.

Session-by-time variables (i.e. PVT-B, CPR quality, T_core_, SOFI) will be assessed for normality of distribution (Shapiro-Wilk test) and analysed with a linear mixed model with session (five levels: CTR, WARM, WARM+FAN, HOT, HOT+FAN) and time (two assessment time points, i.e. pre- and post-fatigue) as fixed effects, and participant as a random effect for repeated measures. Where significant main effects or interactions are identified, pairwise comparisons will be performed between all experimental conditions, with Tukey adjustment for multiple comparisons.

Session variables, including the total nursing performance score and its component behaviours, mean T_skin_, heart rate, and whole-body sweat loss will be analysed using linear mixed models, with session as a fixed effect and participant as a random effect to account for repeated measurements. Pairwise comparisons will be performed between all experimental conditions, with Tukey adjustment for multiple comparisons.

Effect estimates will be reported with 95% confidence intervals. Descriptive statistics will be presented as mean ± standard deviation for approximately normally distributed data, or median and interquartile range for non-normally distributed data. Statistical significance will be accepted at p < 0.05.

### 2.6 Data Management

Data management will be in line with the University of Southampton’ policy on data quality, which forms part of the University’s Information Governance Framework and demonstrates compliance with its obligations under the Data Protection Legislation. Therefore, the study will comply with the requirements of the Data Protection Act 2018 and the University of Southampton’s Ethics Committee (ERGO) policies. This project involves human participants and will be conducted in line with the University’s Policy on the Ethical Conduct of Research and Studies involving Human Participants, and the Medical Research Council’s policies on ethics and data sharing. Data will be fully anonymised at the earliest opportunity and before being made available open access in the University’s data repository. All data that supports publications will be deposited and will be citable using a persistent identifier (DOI). Original hardcopies of study documents (e.g. consent forms) will be stored securely for ten years from completion of the project within a locked office at the University or scanned, encrypted and securely stored on the University’s IT system.

### 2.7 Ethical Considerations and Declarations

The project will involve testing healthy individuals aged 18-65 years and will be conducted in line with Southampton University Code of Practice for Research and will comply with the Declaration of Helsinki. Participants will provide written informed consent, and relevant personal information (e.g. health screen questionnaire). The main risk to participants is heat exposure during low-intensity exercise and simulated nursing work. To minimise this risk, participants will be screened for contraindications to heat exposure, sessions will be time-limited and separated by at least 48 h, and T_core_, T_skin_, heart rate, and environmental conditions will be monitored throughout testing. Trials will be terminated at the participant’s request, at the researcher’s discretion, or if predefined physiological safety thresholds are exceeded. Participants will remain under observation after each session until physiological indicators confirm recovery from heat exposure. Cooling procedures, including fan-assisted cooling and access to cool showers, will be available if required. Minor discomfort may arise from wearable sensors, adhesive tape, ingestion of the telemetric temperature capsule, nude body mass measurement, or being observed during simulated clinical tasks. Participants will be informed in the recruitment materials and participant information sheet that simulated clinical performance data are collected for research purposes only and will not be used as a formal assessment of professional competence. Individual performance data will be anonymised and will not be shared with employers, managers, colleagues, professional bodies, or anyone outside the research team.

Ethical approval for the stated measurements and procedures has been granted by the University of Southampton’s Ethics Committee (ERGO 114314). The study has also been registered prospectively on the ISRCTN registry (https://www.isrctn.com/ISRCTN45793399).

### 2.8 Status and timeline of the study

At the time of publication pilot testing and technical development of the protocol have been completed. Formal recruitment will commence on 1^st^ February 2027. The project will have a lifespan of 24 months.

## 3. Discussion

Heatwaves are becoming increasingly frequent and intense in the UK (3–5, 52). Many NHS hospitals lack adequate cooling infrastructure, leaving healthcare settings vulnerable to indoor overheating during periods of extreme heat (7). As a result, healthcare staff such as nurses may experience increased occupational heat exposure, while attempting to maintain service continuity during periods of heightened operational demand (4, 10, 11). The proposed research aims to characterise the physiological and cognitive burden experienced by hospital nurses working under simulated heatwave conditions and to examine how heat exposure, and its mitigation via cooling, affect their ability to perform routine clinical activities. This is important for advancing scientific understanding of occupational heat strain in healthcare and is timely given the growing need for climate-resilient health systems. The findings may also provide preliminary empirical evidence to inform hospital heatwave plans, implementation of the Adverse Weather and Health Plan (20), and NHS climate adaptation strategies (21) aimed at protecting workforce functioning and patient safety during extreme heat.

### 3.1 Strengths of the planned study

A key strength of the planned study is its use of a controlled climatic chamber to examine the effects of environmental heat exposure on nursing performance under reproducible conditions. Studying healthcare staff during naturally occurring heatwaves is challenging due to variability in staffing levels, workload, environmental conditions, and the practical difficulties of data collection in ward environments. By contrast, the proposed repeated-measures cross-over design will allow each participant to act as their own control, reducing inter-individual variability and enabling direct within-participant comparisons across five thermal conditions: 18°C, 26°C, 34°C, 26°C + fan, and 34°C + fan.

The protocol also combines objective physiological measurements with work-related behavioural and performance-based outcomes during the same simulated clinical shift. T_core_, T_skin_, heart rate, WBSL, fatigue, vigilance, CPR performance, and observational measures of nursing task performance will be collected concurrently. This integrated approach is novel within the healthcare heat-stress literature and will allow the study to characterise both the physiological burden of heat exposure and its potential translation into measurable changes in clinical task performance (12, 23, 53).

Another strength is the applied relevance of the selected environmental conditions. The 34°C exposure reflects high indoor temperatures that have occurred in hospital settings during heatwave conditions, while the 26°C condition serves both as a warm comparison condition, but also as a pragmatic proxy for active cooling (i.e. air conditioning), when compared to the 34°C exposure. Specifically, if ward temperatures exceed recommended or proposed upper limits, i.e. 27°C (22), air-conditioning or other cooling systems may realistically be used to reduce indoor temperatures to approximately 26°C rather than to thermoneutral levels. This condition therefore allows the study to evaluate whether reducing a very hot ward environment to a controlled warm temperature is sufficient to attenuate physiological strain and preserve nursing performance. The fan-assisted cooling conditions will additionally allow the feasibility and potential benefit of a low-cost, rapidly deployable cooling strategy to be evaluated at both 26°C and 34°C. This empirical evidence could inform both hospital heatwave plans, implementation and revisions of the Adverse Weather and Health Plan for England, NHS adaptation policy, and help answering questions for decision makers such as what actions should be triggered (e.g. reducing air temperature vs. providing electric fans) and at what environmental conditions (e.g. air temperature thresholds for concern).

Finally, the simulated nursing ward round, including standardised patient encounters and emergency CPR scenarios, enhances the clinical and operational relevance of the protocol. Although simplified compared with real-world nursing practice, the inclusion of communication, vital-sign assessment, medication management, handover recall, infection-prevention behaviours, and emergency response tasks provides a practical model for examining how heat exposure may affect different components of safety-critical nursing work. By linking these simulated clinical tasks with physiological, fatigue, vigilance, and CPR performance outcomes across defined indoor temperature and fan-assisted cooling conditions, the study may generate preliminary evidence on how heat affects workforce functioning and patient safety risk.

This strengthens the protocol’s relevance to heat-health policy and operational decision-making. In particular, the completed study may help inform hospital heatwave plans, Adverse Weather and Health Plan implementation (20), and NHS climate adaptation strategies (21) by identifying indoor temperature conditions associated with increased concern for staff performance and by evaluating whether fan-assisted cooling is a feasible low-energy mitigation strategy during periods of extreme heat.

### 3.2 Limitations of the planned study

The planned study is not without limitations. First, although the climatic chamber provides strong experimental control, the simulated clinical environment cannot fully reproduce the complexity, unpredictability, and emotional demands of real-world hospital care (25, 54). While the simulated ward round reflects key nursing activities, factors such as competing priorities, interruptions, multidisciplinary communication, patient deterioration, and organisational pressures will necessarily be simplified (55). The inclusion of two cardiac arrest scenarios allows CPR performance to be standardised and assessed, although this frequency of emergency events is unlikely to reflect a typical shift.

Second, the participant sample is restricted to healthy, physically active qualified nurses aged 18-65 years. This is appropriate for a controlled heat-exposure study and reduces safety risks, but may limit generalisability to the wider nursing workforce, including older staff, those with lower physical fitness, health conditions, or medication use that may influence thermoregulation. Findings should therefore be interpreted as mechanistic and feasibility evidence rather than definitive estimates of risk across nursing staff of all characteristics. Yet, it is also important to note that should any adverse physiological or performance effects be observed in our cohort, we expect these effects to be amplified in nurses with greater heat susceptibility, such as those with higher BMI, pre-existing health conditions, or altered thermoregulation.

Third, the repeated-measures design requires participants to complete five experimental sessions, which may introduce learning, familiarisation, or fatigue effects. Condition order will be pseudo-randomised and sessions separated by at least 48 h to minimise these effects; however, some residual learning effects in clinical tasks, cognitive assessments, or CPR performance may remain.

## 4. Conclusion

In conclusion, this protocol describes a controlled, repeated-measures cross-over simulation study designed to determine how environmental heat exposure and electric fan-assisted cooling influence physiological strain and nursing performance during simulated clinical work. By providing a controlled methodological framework for assessing the effects of heat exposure on hospital nurses’ physiological, perceptual, and clinical performance, this study will provide empirical evidence on air temperature-related changes in nursing performance and physiological strain and the role of cooling interventions in modifying these responses. The findings will inform future experimental studies and support the development of evidence-based guidance for hospital heatwave planning, future revisions of the Adverse Weather and Health Plan for England, and NHS climate adaptation strategies aimed at protecting healthcare workers and maintaining patient safety during periods of extreme heat.

## Data Availability

No datasets were generated or analysed during the current study. Data generated from this study will be made publicly available upon study completion.

## Funding

This research is supported by a National Institute for Health and Care Research (NIHR) grant (NIHR506325). The funders did not play any role in the study design, decision to publish or preparation of the manuscript.

## Competing interests

The authors have declared that no competing interests exist.

## Notes

### Competing Interest Statement

The authors have declared no competing interest.

### Clinical Trial

ISRCTN49848

### Author Declarations

Ethical approval for the stated measurements and procedures has been granted by the University of Southampton’s Ethics Committee (ERGO 114314).

